# Antimicrobial Resistance Pattern among Organisms Isolated from Tertiary Care Hospital in Chennai

**DOI:** 10.1101/2025.01.01.25319855

**Authors:** G. F Moses, B Subhasri, R Sakthi Abirami, V Kalpana Devi, U Durga shree, A Rithika, G Bannari Amruth

**Author notes:** **Corresponding Author: Moses. G. F^1^ Address**: No 9/36, Swamy Nagar, Retteri, kolathur, Chennai – 600 099, **Email** –.

## Abstract

**Background:** The findings will help guide the development of targeted antimicrobial stewardship programs and enhance public health strategies to address resistance.

**Aim:** This study aimed to identify common antimicrobial-resistant organisms and their resistance profiles in clinical specimens from a tertiary care hospital.

**Settings and Design:** A cross-sectional study was conducted at ACS Medical College and Hospital, Chennai, from November 2023 to May 2024, involving 346 participants.

**Materials and Methods:** Clinical specimens, including urine, blood, sputum, and wound swabs, were collected from patients in various hospital wards. Demographic details of participants were documented, and patients who had received antimicrobial therapy within the previous month or provided tissue samples were excluded. The specimens were analyzed in the central laboratory for microbial growth and antimicrobial resistance profiles. Isolated were tested by Kirby-Bauer disc diffusion method and breakpoint minimum inhibitory concentration for susceptibility against standard antibiotics.

**Results:** Among the 346 specimens analyzed, 264 (76.3%) showed microbial growth. The most commonly isolated organisms were Escherichia coli (28.8%), Klebsiella species (26.9%). High level resistance was observed, especially to Polymyxin B (98.7%), Cefoperazone/Sulbactam (97.4%), and Cefotaxime (92.1%).

**Conclusion:** antimicrobial resistance requires coordinated action across healthcare, policy, and public sectors. By enforcing stricter regulations, enhancing surveillance, and investing in new treatments, we can slow resistance and preserve the effectiveness of antibiotics, safeguarding public health for future generations.

## INTRODUCTION

Antimicrobial resistance (AMR) presents a critical and escalating threat to global health, significantly undermining the effective treatment of infectious diseases. The World Health Organisation (WHO) identifies AMR as one of the top ten public health challenges worldwide.^[1]^ According to a 2019 report, AMR is linked to approximately 700,000 deaths each year. A figure projected to rise dramatically to 20 million by 2050, with potential economic costs exceeding $2.9 trillion.^[2]^

In India, the situation regarding AMR is particularly alarming due to several interconnected factors:

- **Over-the-counter Antibiotics:** Antibiotics are readily accessible without prescriptions, facilitating misuse and self-medication.
- **Weak Regulatory Framework:** Insufficient regulatory measures and regulations on antibiotic use.
- **Lack of Awareness:** Understanding of AMR in India is low, many people do not understand the implications of not finishing a course of antibiotic or using them without necessity, Which can lead to bacteria evolving resistance against these medications.^[3]^

Tertiary care hospitals, which handle a high volume of critically ill patients, often become hotspots for resistant pathogens. The emergence of bacterial resistance is further exacerbated by:

- **Antibiotic Misuse:** Inappropriate or unnecessary use of antibiotics by patients and healthcare providers alike.
- **Misdiagnosis:** Errors in diagnosing infections can lead to inappropriate treatment choices.
- **Substandard Prescribing Practices:** Some prescribers do not consistently adhere to best practices for antibiotic use.
- **Patient Self-Medication:** Individuals frequently take antibiotics without professional oversight, exacerbating resistance.
- **Inadequate Healthcare Conditions:** Poor hygiene and insufficient healthcare infrastructure facilitate the spread of resistant bacteria.
- **Agricultural Antibiotic Use:** Excessive use of antibiotics in agriculture significantly contributes to the resistance problem.

This study aims to investigate the prevalence and patterns of AMR among clinical isolates at a tertiary care hospital in Chennai. Our specific objectives are to:

1. Identify the most common resistant organisms.
2. Propose effective strategies for antimicrobial stewardship.

This research underscores the urgent need for collective action and provides a comprehensive roadmap to tackle the AMR challenge. By creating awareness and collaboration among healthcare providers, policymakers, and the community, we can effectively combat AMR and ensure the continued efficacy of life-saving antibiotics for future generations.

## MATERIALS AND METHODS

This cross-sectional study was conducted from November 2023 to May 2024 at ACS Medical College and Hospital in Chennai, Tamil Nadu, following approval from the institutional ethics committee (No. 975/2023/IEC/ACSMCH Dt. 17.11.2023).

### Study design

cross sectional study

The hospital encompasses various departments, including:

1. General Medicine
2. General Surgery
3. Orthopaedics
4. Gynaecology and Obstetrics
5. Pediatrics
6. Pulmonology
7. Dermatology

This diverse clinical environment facilitates robust research opportunities. A total of 346 participants were enrolled in the study.

### Inclusion Criteria

1. **Specimens:**

a. Urine
b. Blood
c. Sputum
d. Wound/Pus
e. Additional samples (ear swabs, vaginal swabs)

Processing: All specimens processed under aseptic conditions to ensure minimal contamination.

### Exclusion Criteria

1. **Tissue Samples**: Any tissue samples were excluded from analysis.
2. **Prior Antimicrobial Therapy**: Individuals who had received any form of antimicrobial therapy prior to specimen collection were excluded to maintain the integrity of the findings.

This structured approach ensured a reliable analysis of microbial cultures from the selected specimens. Specimens were sent to the central laboratory for comprehensive culture analysis. Staining and biochemical tests were done for preliminary identification of the isolates.

#### Antibiotic sensitivity testing

Kirby-Bauer’s disk diffusion method on Muller Hinton agar were used for antibiotic sensitivity testing and interpreted based on Clinical and Laboratory Standards Institute (CLSI) guidelines^[4]^

The results were compiled into an antibiogram (An antibiogram is a laboratory report that shows the susceptibility of bacterial strains to various antibiotics, helping guide effective treatment options), summarising the susceptibility patterns of the isolated organisms. Data were gathered from the laboratory’s antibiotic sensitivity testing reports.

Statistical analysis was performed using SPSS version 2.0. Frequencies and percentages were calculated for demographic and clinical variables. The chi-square test was used to compare categorical variables, while the Mann-Whitney U test was applied to analyse continuous variables. Fisher’s exact test was utilized for small sample sizes. This approach ensures clarity and reproducibility in the data analysis.

## Results

The study analysed 346 samples, revealing significant demographic insights that can inform our understanding of antimicrobial resistance.

### Age Distribution

The predominance of older adults (56.6%) highlights their increased vulnerability to infections due to age-related health challenges and a less effective immune system. This population often requires longer hospital stays and more aggressive treatment approaches, including the use of broad-spectrum antibiotics, which can increase exposure to resistant organisms, particularly Gram-negative bacteria (75.4%).

### Gender Distribution

The sample included a slightly higher proportion of females (52.3%) compared to males (47.7%). Significant differences in infection rates or resistance patterns may arise from biological and behavioural factors related to gender.

### Sample Type Distribution

Urine samples constituted the largest portion (42.4%), reflecting the common occurrence of urinary tract infections, particularly in older adults. The significant number of wound/pus and sputum/swab samples (24.8% each) suggests a range of infections that could be influenced by prior antibiotic use. The lower percentage of blood samples (5.2%) highlights the need for vigilance in systemic infections (table 1).

**Table 1:** Demographic Profile enrolled in this study from November 2023 to May 2024.

| Distribution of Age |  |  |
| --- | --- | --- |
| Age | Frequency(n=346) | Percentage |
| Old age adults (above 45) | 196 | 56.6% |
| Middle-aged adults (31-45) | 77 | 22.1% |
| Young adults (17-30) | 45 | 13% |
| Child (0-16) | 28 | 8% |
| Distribution of Gender |  |  |
| Gender | Frequency(n=346) | Percentage |
| Female | 181 | 52.3% |
| Male | 165 | 47.7% |
| Distribution of Sample Type |  |  |
| Sample type | Frequency(n=346) | Percentage |
| Urine | 147 | 42.4% |
| Wound/pus | 86 | 24.8% |
| Sputum/swab | 86 | 24.8% |
| Blood | 18 | 5.2% |
| Fluid, ear swab | 9 | 2.6% |

### Overall Culture Results

Out of 346 samples analysed, 264 (76.3%) exhibited positive growth in cultures, while 82 samples (23.7%) showed no growth.

### Prevalence of Gram-negative Bacteria

Gram-negative organisms accounted for 75.4% of the positive cultures, highlighting their dominance in the infections present within this population. This prevalence is concerning, as Gram-negative bacteria are often associated with increased antibiotic resistance.

### Sample Type Analysis

- **Urine**: A significant proportion of Gram-negative bacteria (48.7%) were isolated from urine samples, aligning with the common occurrence of urinary tract infections, particularly in older adults.
- **Wound/Pus**: This sample type showed a notable prevalence of Gram-positive bacteria (55.5%), indicating that skin and soft tissue infections may be more associated with these organisms. This distinction suggests a need for targeted therapies based on sample origin.
- **Sputum**: Both Gram-negative (24.6%) and Gram-positive (24.6%) organisms were similarly represented in sputum samples, indicating a diverse range of potential pathogens in respiratory infections.
- **Blood**: The low percentages for blood samples (3.5% Gram-negative and 6.1% Gram-positive) underscore the importance of monitoring systemic infections, even if they are less frequent (table 2).

**Table 2:** Distribution of organisms grown, and analysis of culture type with Gram Stain.

| Distribution of Organisms Growth |  |  |  |  |
| --- | --- | --- | --- | --- |
| Culture | Frequency(n=346) |  | Percentage |  |
| Positive | 264 |  | 76.3% |  |
| No growth | 82 |  | 23.7% |  |
| Distribution of sample type between Gram negative and Gram positive |  |  |  |  |
| Culture | Gram Negative |  | Gram Positive |  |
|  | Frequency<br>(n)=199 | Percentage (%) | Frequency<br>(n)=65 | Percentage<br>(%) |
| Urine | 97 | 48.7% | 9 | 13.8% |
| Wound/pus | 43 | 21.6% | 36 | 55.5% |
| Sputum | 49 | 24.6% | 16 | 24.6% |
| Blood | 7 | 3.5% | 4 | 6.1% |
| Others | 3 | 1.5% | - | - |
| Total | 199 | 75.4% | 65 | 24.6% |

Of the 264 positive cultures, 40.2% were from urine samples, 29.9% from wound/pus samples, 24.6% from sputum samples, 4.2% from blood samples, and 1.1% from other specimens such as ear and vaginal swabs (table 3).

**Table 3:** Distribution of Sample Type for shown positive growth.

| Sample type | Frequency (n=264) | Percentage |
| --- | --- | --- |
| Urine | 106 | 40.2% |
| Wound/Pus | 79 | 29.9% |
| Sputum | 65 | 24.6% |
| Blood | 11 | 4.2% |
| Others | 3 | 1.1% |

### Prevalence of E. coli

E. coli is the most frequently isolated organism, particularly from urine samples (51.9%), which is consistent with the high incidence of urinary tract infections in older adults. Its significant presence in other sample types indicates its versatility as a pathogen.

### Klebsiella spp.

This organism shows a substantial presence across various samples, especially in sputum (43.1%) and wound cultures (17.7%). Its prevalence in respiratory infections raises concerns about its potential for multi-drug resistance, especially in hospitalised patients.

### Staphylococcus spp.

Notably higher in wound samples (30.4%), this genus includes both pathogenic and non-pathogenic species. Its presence suggests skin and soft tissue infections, which may require careful antibiotic selection to address resistance.

### Pseudomonas spp.

Found in both wound (8.9%) and sputum (18.5%) samples, Pseudomonas is known for its antibiotic resistance, making it a significant concern in healthcare settings.

### Salmonella spp.

Detected exclusively in blood samples (36.3%), this organism highlights the need for prompt identification and treatment, as systemic infections can be life-threatening.

### Diverse Organism Presence

The variety of organisms isolated from different sample types emphasises the complexity of infections in the study population. It suggests a need for tailored treatment approaches based on the specific pathogens identified in each clinical context (table 4).

**Table 4:**
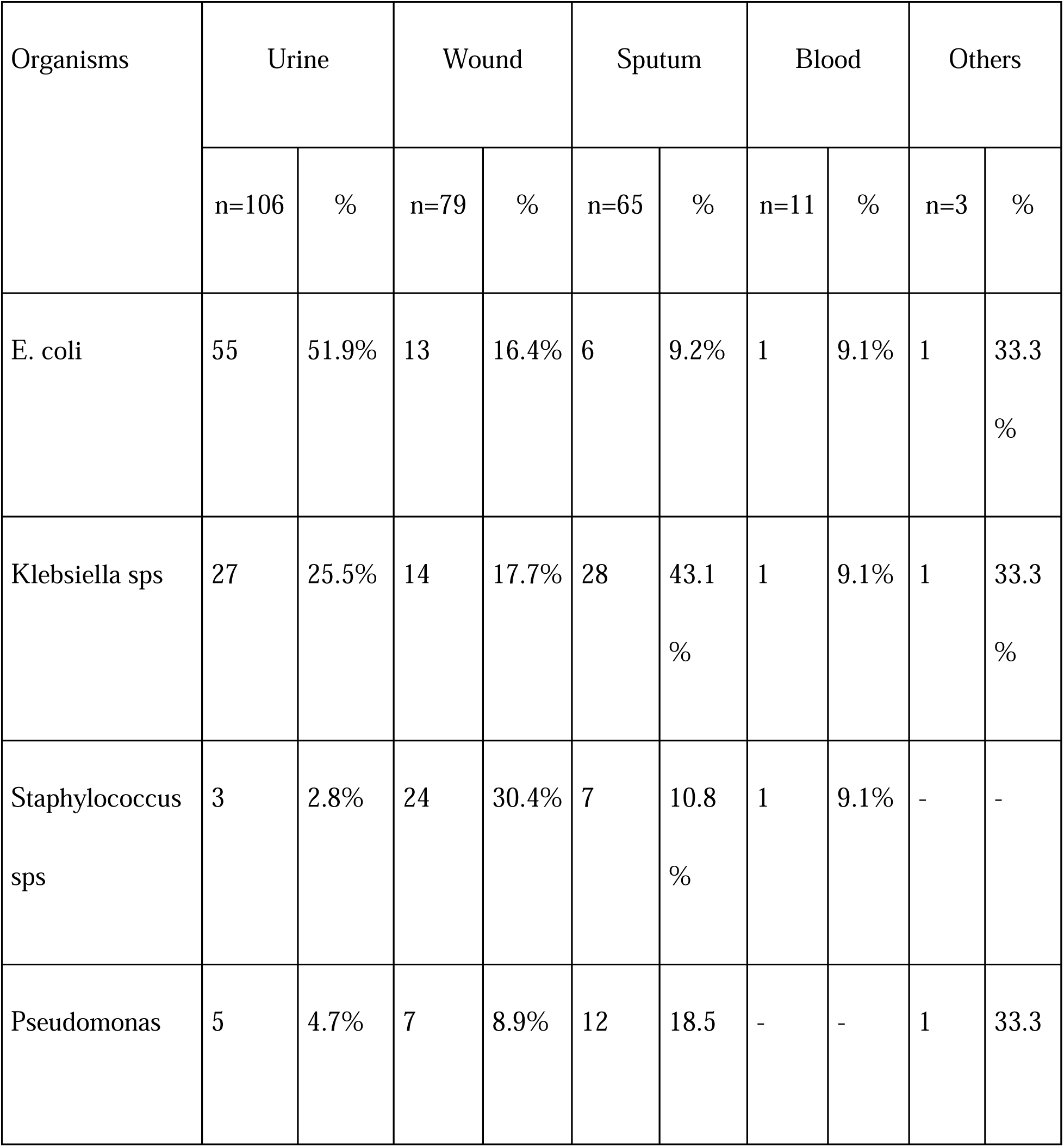
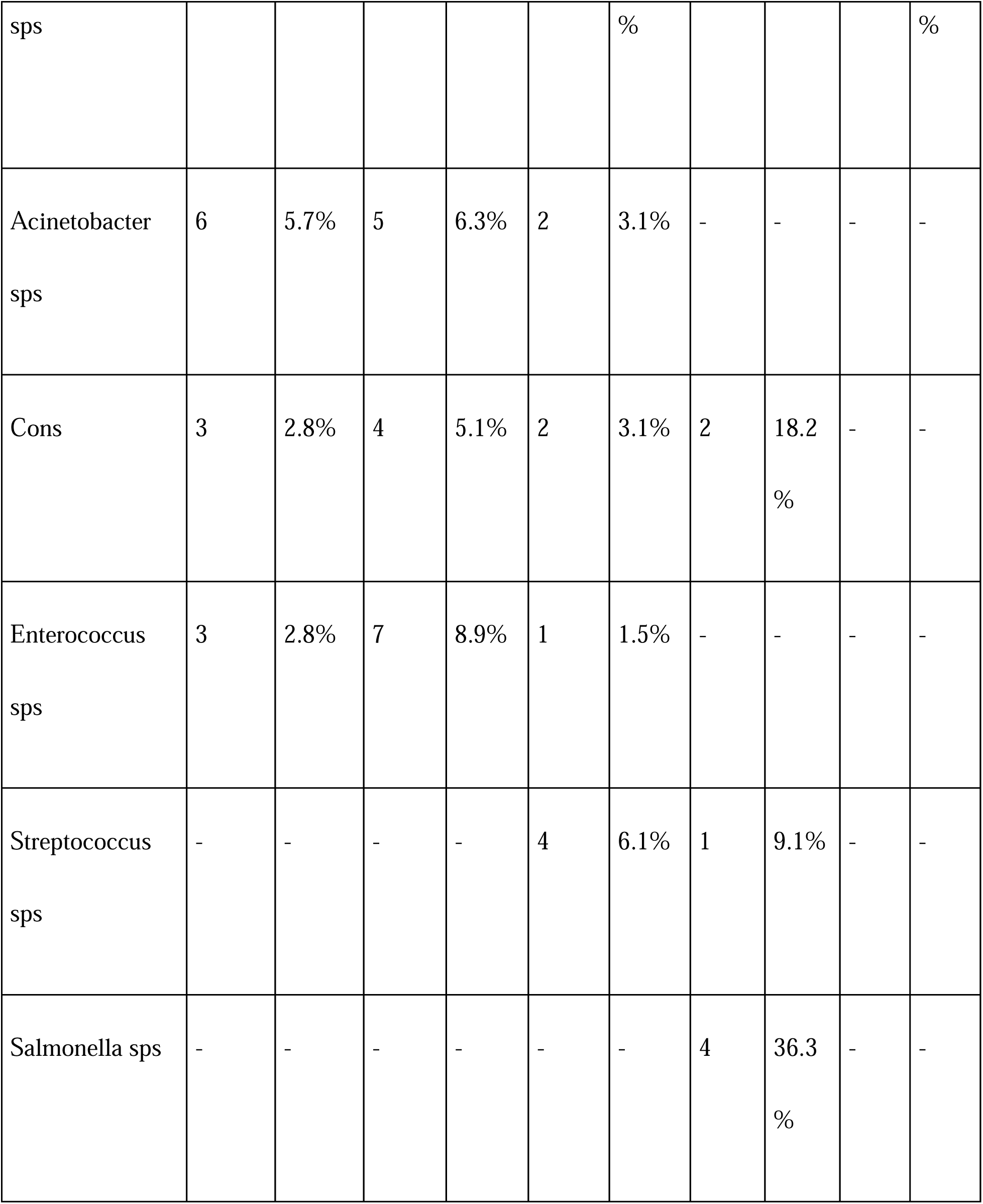
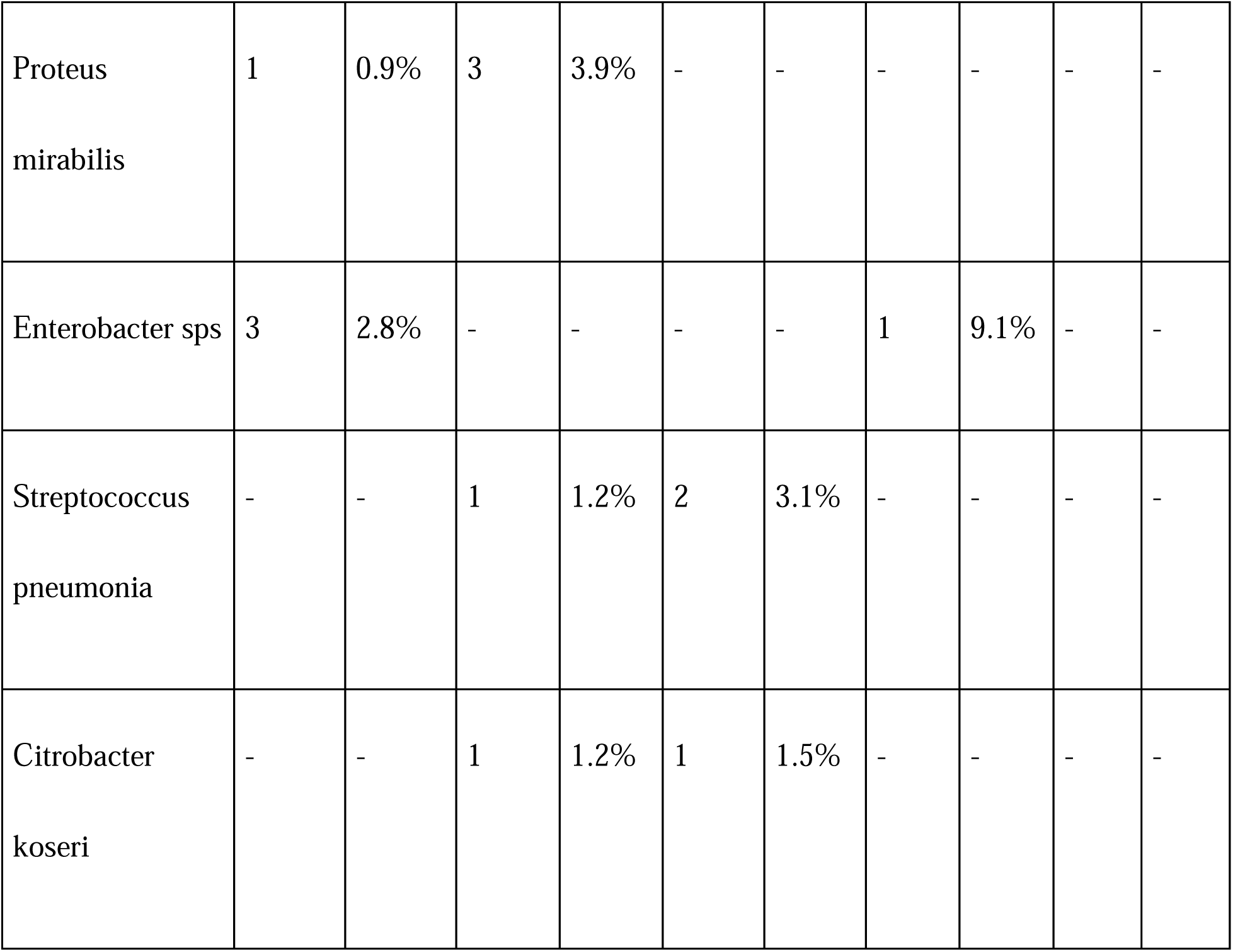
Analysis of Organisms with sample types.

1. **Dominance of E. coli:** E. coli accounted for 28.8% of all positive cultures, reaffirming its status as a leading pathogen across multiple infection types. The high prevalence in urine samples suggests its central role in urinary tract infections.
2. **Prevalence of Klebsiella spp.:** Overall, Klebsiella spp. constituted 26.9% of positive cultures. Its prominence in both sputum and urine emphasises its role in respiratory and urinary infections, raising concerns about resistance patterns.
3. **Staphylococcus spp. Distribution:** Staphylococcus spp. made up 13.2% of all positive cultures, showing that while it is particularly significant in wound infections, it is also a notable pathogen in other infection types.
4. **Presence of Pseudomonas spp.:** Overall, Pseudomonas spp. accounted for 9.5% of positive cultures, consistent with its classification as an opportunistic pathogen that poses a risk in healthcare settings.
5. **Other Organisms:** The overall representation of these organisms is lower (4.9% for Acinetobacter spp. and 4.2% for Enterococcus spp.); indicating that while they are present, they may not be as prevalent as E. coli or Klebsiella spp. across all samples (table 5).

**Table 5:**
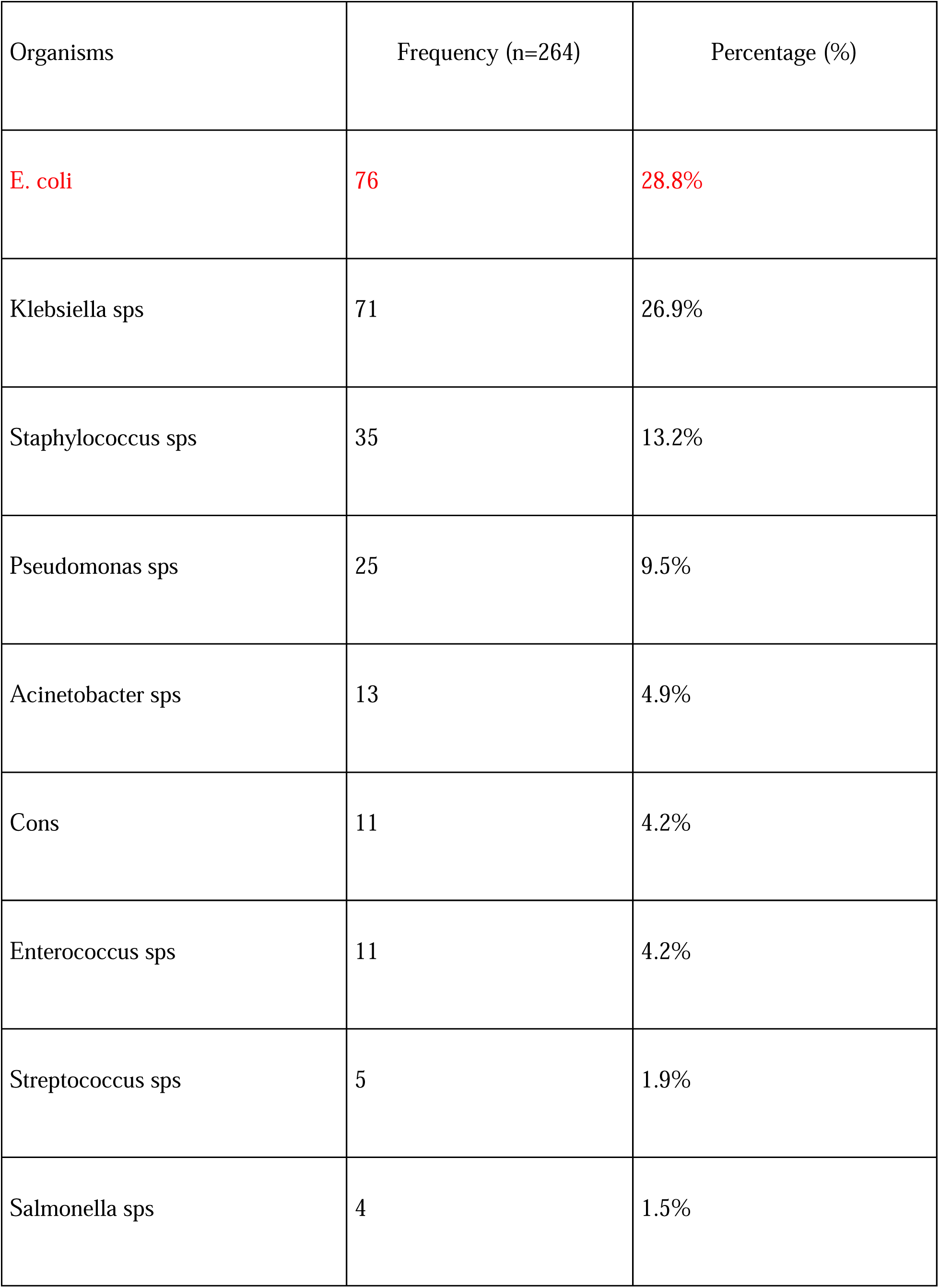
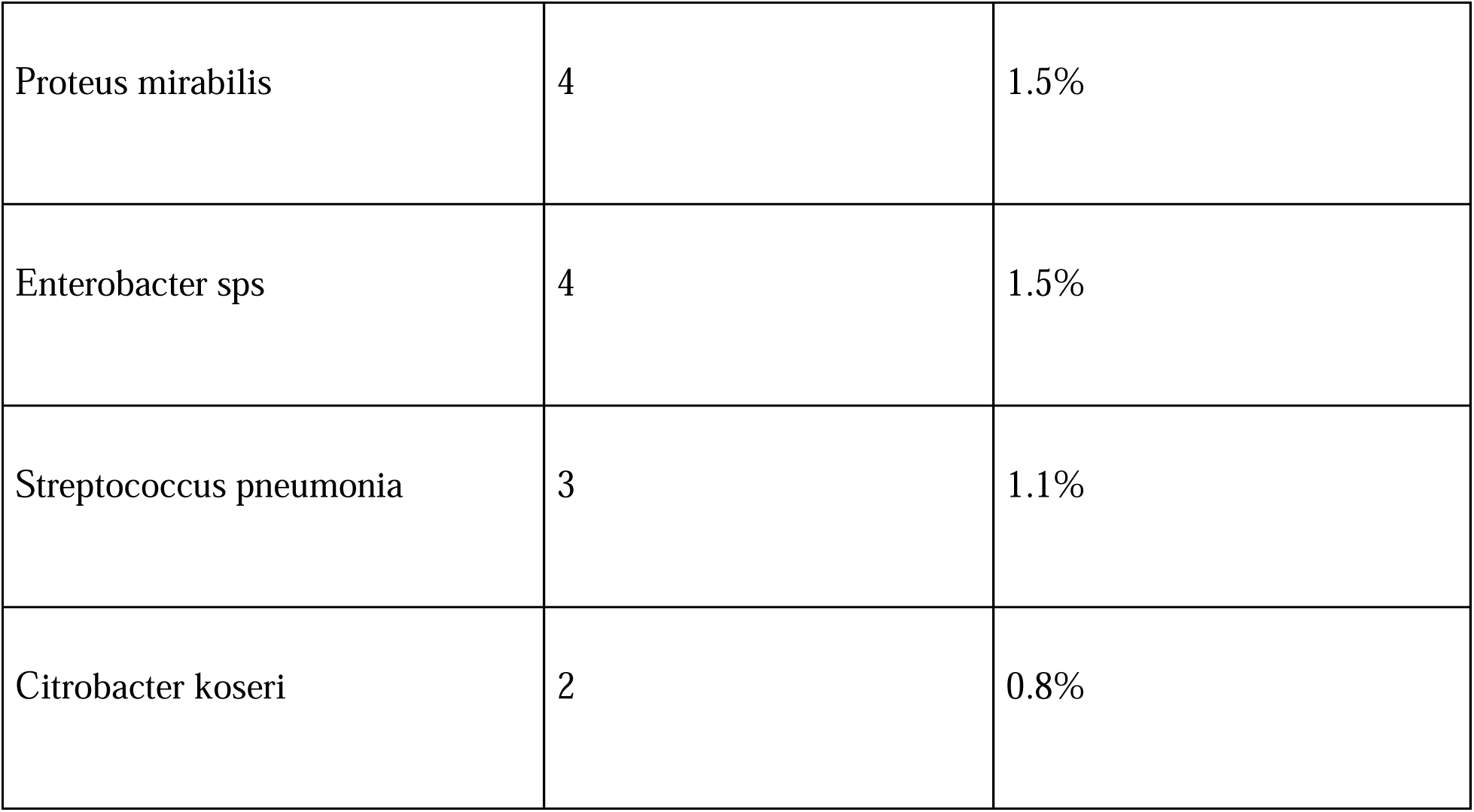
Various Organisms that shown Positive culture.

A notable percentage of resistance is observed for several antibiotics, particularly: **Cefoperazone /sulbactam** (97.4%), **Cefotaxime** (92.1%), **Cefuroxime** (93.4%), and **Polymyxin B** (98.7%). These high resistance rates among third-generation cephalosporins and Polymyxin B indicate a pressing challenge in treating infections caused by bacteria sensitive to these antibiotics.

### Effective Options

Some antibiotics exhibit a higher sensitivity rate, including **Amikacin** (71.1%) and **Gentamycin** (63.2%), suggesting they may be more effective choices in treating infections caused by the tested bacteria.

### Intermediate Responses

Several antibiotics show significant intermediate responses: **Ampicillin** (30.3%), **Ceftriaxone** (50%), **Levofloxacin** (15.8%), and **Co-trimoxazole** (38.2%). While not outright resistant, careful consideration is needed when using these drugs, as their effectiveness may vary (table 6).

**Table 6:** Resistance pattern of E. coli (n=76)

| Antibiotics | Sensitive |  | Intermediate |  | Resistance |  |
| --- | --- | --- | --- | --- | --- | --- |
|  | Frequency | Percentage | Frequency | Percentage | Frequency | Percentage |
| Ampicillin | 9 | 11.8% | 44 | 57.9% | 23 | 30.3% |
| Gentamycin | 48 | 63.2% | 14 | 18.4% | 14 | 18.4% |
| Amikacin | 54 | 71.1% | 8 | 10.5% | 14 | 18.4% |
| Amoxiclav | 14 | 18.4% | 18 | 23.7% | 44 | 57.9% |
| Piperacillin/<br>tazobactam | 44 | 57.9% | 13 | 17.1% | 19 | 25% |
| Cefeperazone/<br>sulbactam | 1 | 1.3% | 1 | 1.3% | 74 | 97.4% |
| Cefotaxime | 1 | 1.3% | 5 | 6.6% | 70 | 92.1% |
| Ceftriaxone | 20 | 26.3% | 38 | 50% | 18 | 23.7% |
| Ciprofloxacin | 27 | 35.5% | 33 | 43.4% | 16 | 21.1% |
| Co-trimoxazole | 32 | 42.1% | 29 | 38.2% | 15 | 19.7% |
| Cefazolin | 10 | 13.2% | 35 | 46% | 31 | 40.8% |
| Levofloxacin | 11 | 4.5% | 12 | 15.8% | 53 | 69.7% |
| Ceftazidime | 36 | 47.4% | 12 | 15.8% | 28 | 36.8% |
| Ceftazidime with<br>clav | 1 | 1.3% | - | - | 75 | 98.7% |
| Chloramphenicol | 2 | 2.6% | - | - | 74 | 97.4% |
| Tetracycline | 30 | 39.5% | 27 | 35.5% | 19 | 25% |
| Cefuroxime | 4 | 5.3% | 1 | 1.3% | 71 | 93.4% |
| Cefepime | 33 | 43.4% | 20 | 26.3% | 23 | 30.3% |
| Polymyxin –b | - | - | 1 | 1.3% | 75 | 98.7% |
| Vancomycin | - | - | 1 | 1.3% | 75 | 98.7% |
| Linezolid | 1 | 1.3% | - | - | 75 | 98.7% |
| Azithromycin | 19 | 25% | 20 | 26.3% | 37 | 48.7% |
| Imipenem | 8 | 10.5% | 46 | 60.5% | 22 | 29% |
| Meropenem | 6 | 7.9% | 46 | 60.5% | 24 | 31.6% |

## Discussion

### Principal Findings

This study examined antimicrobial resistance (AMR) patterns in 346 patients. The key finding is the dominance of Gram-negative bacteria, which accounted for 75.4% of positive cultures. Among these, Escherichia coli and Klebsiella species were the most frequently isolated organisms. The high prevalence of E. coli in urinary tract infections (UTIs) is significant. Klebsiella species were found in both sputum and urine samples, with a higher prevalence in sputum and being the second most common pathogen in urine samples. This underscores Klebsiella’s important role in both respiratory and urinary tract infections. Infections caused by various organisms are particularly prominent among older adults, as they are more prone to infections due to age-related factors. Additionally, resistance to several important antibiotics was observed, particularly third-generation cephalosporins and Polymyxin B. These high resistance rates are concerning, indicating a growing challenge of multi-drug resistance.

### Key Findings and Strengths of the Study

- **Pathogen Identification:** The study highlights common pathogens like E. coli, Klebsiella, and Pseudomonas, along with their resistance profiles, aiding in infection source identification.
- **Resistance Data:** Comprehensive data on multidrug resistance in both Gram-negative and Gram-positive bacteria is provided, essential for antimicrobial stewardship programs.
- **Targeted Stewardship:** High resistance rates for specific pathogens and antibiotics the development of locally tailored stewardship strategies to improve patient outcomes.
- **Sample Diversity:** Detailed infection site breakdowns (urine, wound, sputum, blood) offer insights into pathogen distribution.
- **Age Distribution:** The study emphasizes the increased vulnerability of older adults to infections, highlighting age as a key factor in susceptibility.
- **Resistance Patterns:** Detailed profiles, especially against third-generation cephalosporins and polymyxins, emphasize significant resistance challenges.

### Limitations of the Study

- **Short Duration:** The six-month timeframe limits the ability to capture seasonal variations and long-term AMR trends.
- **Small Sample Size:** A low sample size may affect the accuracy and generalizability of the results.
- **Rapid AMR Changes:** AMR patterns may evolve quickly, making the findings less relevant over time.
- **Cross-Sectional Design:** The study’s design restricts the ability to assess trends or causal relationships.
- **Context-Specific:** Results may not be applicable to other hospitals or regions with different resistance profiles.
- **Rural Focus:** The study’s focus on urban settings limits its applicability to rural healthcare contexts, where AMR patterns may differ, as highlighted by Mardourian et al. (2013). Including rural or underserved areas could provide a more comprehensive understanding of AMR in these regions.^[5]^
- **Missing Clinical Data:** The study lacked clinical outcome and co-morbidity data, which are essential for understanding resistance patterns, as highlighted in Gupta et al. (2001).^[6]^
- **Non-Hospital-Acquired Infections:** The study did not focus on hospital-acquired infections, which are important for understanding resistance in healthcare settings, as noted in Saravanan and Raveendaran (2013).^[7]^

### Discussion of Important Differences in Results

One of the most striking findings in our study is the exceptionally high resistance rates observed for several antibiotics. Resistance to third-generation cephalosporins, such as Cefotaxime (92.1%) and Cefuroxime (93.4%), was notably high, which may be attributed to widespread overuse of these antibiotics in hospital settings, particularly for empirical therapy. The near-complete resistance observed for Polymyxin B (98.7%) is especially concerning, as it is often reserved as a last-line treatment for multidrug-resistant organisms. This may indicate the emergence of super-resistant strains that pose significant treatment challenges. These findings highlight the need for stricter antimicrobial stewardship practices, especially in high-risk populations like older adults who are more prone to infections and prolonged antibiotic courses.

In contrast, Amikacin and Gentamicin demonstrated relatively higher sensitivity rates (71.1 % and 63.2%, respectively), suggesting that these antibiotics may offer more effective treatment options for some resistant infections. However, the presence of intermediate responses to many antibiotics, including Levofloxacin (69.7% resistant), Ciprofloxacin (21.1% resistant), and Ceftriaxone (23.7% resistant), indicates a need for careful consideration of local resistance patterns before prescribing.

### Recommendations for Antimicrobial Stewardship

1. **Establish a Multidisciplinary Team:** Form a team of infectious disease specialists, pharmacists, and clinicians to lead stewardship efforts.
2. **Regular Resistance Surveillance:** Monitor antimicrobial resistance trends to guide prescribing practices.
3. **Ongoing Education:** Provide continuous training for healthcare providers on antibiotic use and resistance.
4. **Evidence-Based Guidelines:** Develop and disseminate treatment guidelines based on local resistance data.
5. **Promote Diagnostic Testing:** Utilise rapid tests to confirm infections before initiating antibiotic therapy.
6. **Control Over-the-Counter Antibiotic Sales:** Implement regulations to prevent pharmacies from selling antibiotics without prescriptions.
7. **Control Self-Prescribing Practices:** Address the issue of individuals taking antibiotics without medical guidance to ensure proper prescriptions.
8. **Decision-Support Tools:** Integrate clinical decision-support systems in electronic health records.
9. **Cultivate Accountability:** Encourage open discussions on antibiotic prescribing among clinicians.
10. **Monitor and Adapt:** Regularly assess stewardship outcomes and adjust strategies as needed.

### Future Research Directions

#### Resistance Mechanisms

Study the genetic factors behind resistance to guide better diagnostics and treatments.

#### Antibiotic Stewardship

Evaluate the long-term effectiveness of antibiotic stewardship in reducing resistance, particularly in high-risk groups like the elderly.

#### Host Factors and Microbiome

Investigate how the immune system and microbiome influence susceptibility to resistant infections in older adults.

#### Alternative Therapies

Explore alternative treatments like bacteriophage therapy for multi-drug resistant bacteria.

#### Surveillance Systems

Improve surveillance to track resistance trends and inform infection control and treatment strategies.

## Data Availability

All data produced in the present work are contained in the manuscript

